# Association of Gensini score with inflammatory-metabolic markers in women with CAD: a multidimensional analysis

**DOI:** 10.1101/2025.09.17.25335822

**Authors:** An GuoXia, Liu XiaoHua, Li Ruizhe, Guo Weimin

## Abstract

**Aims:** The diagnosis and assessment of coronary artery disease (CAD) in women remain challenging, as conventional diagnostic tools often show limited sensitivity and risk underdiagnosis. This study aimed to examine the associations between routinely available clinical and laboratory parameters and the severity of coronary artery lesions, quantified by the Gensini score, in female patients. In addition, we sought to develop and validate a predictive model to identify women at high risk of severe atherosclerosis.

**Methods:** This retrospective, cross-sectional study was conducted at the Second Hospital of Shanxi Medical University and included 91 patients with established CAD. Laboratory assessments comprised complete blood counts, high-sensitivity C-reactive protein (hs-CRP), interleukin-6 (IL-6), lipid profiles, and uric acid (UA). Sleep quality was measured using the Athens Insomnia Scale (AIS). Statistical analyses included Spearman correlation, multiple linear regression, and logistic regression to evaluate the relationships between these variables and the Gensini score, and to assess their predictive value for high-risk disease.

**Results:** In univariate analysis, the Gensini score was significantly correlated with inflammatory markers (IL-6: r = 0.61, P < 0.001; hs-CRP: r = 0.43, P = 0.012) and diabetes (r = 0.36, P = 0.037). Multiple linear regression identified insomnia, hypertension, diabetes, IL-6, and hs-CRP as independent predictors of the Gensini score, yielding a high goodness of fit (R² = 0.823, adjusted R² = 0.783, F = 20.19, P < 0.001). A logistic regression model for predicting high-risk disease (Gensini score >20) demonstrated excellent discrimination, with an area under the curve (AUC) of 0.964 in the training set and 0.833 in the test set.

**Conclusion:** In women with CAD, the severity of coronary atherosclerosis is strongly associated with inflammation, metabolic dysregulation, and insomnia, with risk profiles distinct from those observed in men. A significant interaction between hs-CRP and diabetes was identified as a key modulator of CAD severity. These findings highlight the value of integrated statistical models that account for interacting risk factors, offering improved risk stratification and supporting proactive, personalized interventions in female patients with CAD.

## 1. Introduction

A significant paradigm shift is emerging in coronary heart disease (CHD) research. Traditionally, investigations have centered on ischemic symptoms and acute cardiovascular events. Increasingly, however, attention is turning to atherosclerosis—the underlying pathological process of CHD. A landmark publication in *The Lancet* has called for a redefinition of CHD as Atherosclerotic Coronary Artery Disease (ACAD) [1]. This reclassification seeks to redirect clinical research and practice from late-stage interventions toward the early prevention, detection, and management of atherosclerosis, ultimately aiming to reduce the global disease burden. Atherosclerosis is a lifelong process that begins in childhood, characterized by endothelial dysfunction, lipid accumulation, and inflammatory activation. Its progression is driven by a combination of traditional and emerging risk factors.

Among emerging risk factors, the role of sleep health has gained increasing recognition. In 2022, the American Heart Association (AHA) incorporated sleep duration into its *Life’s Essential Eight*, a major update to the *Life’s Simple Seven* proposed in 2010. Sleep duration has since been identified as an important determinant of Coronary Artery Disease (CAD) severity. A study in a Chinese cohort demonstrated that a sleep duration of less than six hours is significantly associated with a higher Gensini score [2], a comprehensive index for quantifying atherosclerotic burden using coronary angiography [3]. In addition, multiple meta-analyses have consistently reported that short sleep duration (≤5 or <6 hours) constitutes a major risk factor for myocardial infarction (MI) and cardiovascular disease.

Insomnia, in particular, has been strongly linked to outcomes such as MI (RR ≈ 1.69), CHD, and stroke [4]. Given that atherosclerosis represents the underlying pathology of these endpoints, disease processes driven by sleep disturbances are inevitably reflected in anatomical scoring systems that measure atherosclerotic burden, such as the Gensini score.

At the same time, recognition of the central role of inflammation in atherosclerosis progression has spurred the identification of novel biomarkers. The Systemic Immune-Inflammation Response Index (SIIRI), first reported by Mangales et al. in 2024, has been shown to predict the severity of coronary artery lesions in patients with acute coronary syndrome [5]. Multivariate analysis in their study confirmed SIIRI as an independent predictor of severe CAD.

Despite these advances, limited research has explored the combined impact of insomnia and systemic inflammation on CAD severity, especially in female patients. To address this gap, our present study investigates the risk factors influencing the onset and progression of CAD in women through multidimensional correlation and regression analyses. By leveraging accessible clinical examinations, laboratory indices, and validated scales, we aim to provide a practical reference for assessing CAD severity. This may, in turn, inform decisions regarding pharmacological or interventional treatment, thereby preventing serious clinical events and optimizing the allocation of medical resources.

## 2. Materials and Methods

### 2.1. Study Design and Population

This cross-sectional study was conducted from October to December 2024 at the Second Hospital of Shanxi Medical University. A total of 91 inpatients with a diagnosis of coronary heart disease (CHD) confirmed by coronary angiography were consecutively enrolled, including 58 males and 33 females.

### 2.2. Exclusion Criteria

Patients were excluded if they met any of the following criteria:

1. autoimmune diseases, severe systemic illnesses, a history of malignancy and/or prior chemotherapy, evidence of concomitant inflammatory diseases, acute infections, chronic inflammatory conditions, other acute cardiovascular diseases, or significant hepatic or renal dysfunction;
2. glucocorticoid therapy within the past 3 months, secondary hypertension, heart failure, or a history of cerebrovascular disease;
3. dependence on or abuse of alcohol, tobacco, or other substances;
4. psychiatric disorders including depression, schizophrenia, or other major mental illnesses.

### 2.3. Data Collection and Clinical Assessments

#### Baseline Characteristics

Demographic data (sex, age), lifestyle habits (smoking and alcohol consumption), medication history, and past medical history were collected using a standardized questionnaire. Diagnoses of CHD, hypertension, diabetes mellitus, hyperlipidemia, and hyperuricemia were made according to current clinical guidelines and consensus statements. Physical examinations included measurements of blood pressure, heart rate, height, and weight.

#### Insomnia Assessment

Upon admission, insomnia severity was evaluated by experienced physicians using the Athens Insomnia Scale (AIS) [6]. Scores were classified as follows: 0–3, normal sleep; 4–5, possible sleep disturbance; and ≥6, insomnia requiring intervention.

### 2.4. Laboratory Measurements

Morning fasting venous blood samples were collected after at least 12 hours of overnight fasting. Laboratory analyses included complete blood count, biochemical profiles, coagulation function, lipid profiles, glycated hemoglobin A1c (HbA1c), high-sensitivity C-reactive protein (hs-CRP), interleukin-6 (IL-6), uric acid (UA), and CD3+ T-cell percentage.

The Systemic Immune-Inflammation Response Index (SIIRI) was calculated using the following formula:

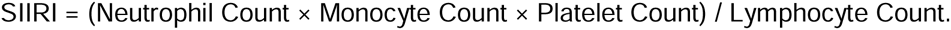

### 2.5. Coronary Angiography and Gensini Score Assessment

All patients underwent coronary angiography (CAG). Coronary artery disease was defined as ≥50% stenosis in at least one major coronary artery. The severity of coronary stenosis was quantified using the Gensini scoring system [3], which incorporates three components: the degree of luminal narrowing, a location-based weighting factor, and a collateral adjustment factor.

- **Severity score:** 1 (<25% stenosis), 2 (25–50%), 4 (51–75%), 8 (76–90%), 16 (91–99%), and 32 (total occlusion, ≥99%).
- **Location multiplier:** left main coronary artery, ×5.0; proximal left anterior descending (LAD) artery, ×2.5; mid-LAD, ×1.5; distal LAD, ×1.0; first diagonal branch (D1), ×1.0; second diagonal branch (D2), ×0.5; proximal left circumflex (LCx), ×2.5; distal LCx and posterolateral branches, ×1.0; obtuse marginal branches, ×0.5; right coronary artery (proximal, mid, distal segments) and posterior descending artery, ×1.0.

The Gensini score for each lesion was calculated as the product of the severity score and the location multiplier. The total Gensini score was obtained by summing the scores of all lesions for each patient.

### 2.6 Statistical analysis

All data analyses were performed in the Python environment (v3.11.9). First, raw data were preprocessed: missing values of numerical variables were imputed using the median, whereas categorical variables were imputed using the mode.

Exploratory data analysis (EDA) and correlation assessments were conducted using the *pandas* (v1.5.3) and *scipy* (v1.13.1) packages. We specifically evaluated the associations between the Gensini score and multiple risk factors, including hypertension, diabetes mellitus, high-sensitivity C-reactive protein (hs-CRP), interleukin-6 (IL-6), Athens Insomnia Scale (AIS), low-density lipoprotein cholesterol (LDL-C), triglycerides (TG), and uric acid (UA).

To further quantify the relationship between these factors and the Gensini score, we constructed a multivariable linear regression model using the *statsmodels* package (v0.14.2). In addition, to assess the predictive value of key risk factors for the severity of coronary artery disease (dichotomized according to the Gensini score), logistic regression analyses were performed with the *scikit-learn* package (v1.5.1).

All data visualizations, including the generation of figures and plots, were carried out using *matplotlib* (v3.10.6) and *seaborn* (v0.13.2). A two-sided *P* value <0.05 was considered statistically significant in all analyses.

### 2.7 Ethical Considerations

This study was approved by the Institutional Review Board of Second Hospital of Shanxi Medical University. As the study employed a retrospective design using medical records, the requirement for informed consent was waived by the IRB. All data were anonymized prior to analysis. Patient confidentiality was strictly maintained, with only the principal investigator, supervisor, and data collectors having access to the data. The study was conducted in accordance with the ethical principles outlined in the Declaration of Helsinki.

## 3. Results

### 3.1 Demographics and Baseline Characteristics

Among the 33 female patients with CHD, the mean age was 61.52 years, and the median Gensini score was 24. In contrast, the 58 male patients had a mean age of 57.26 years and a median Gensini score of 30. Normally distributed variables are expressed as mean ± standard deviation (SD), whereas non-normally distributed variables are expressed as median values.

**Table 1.**
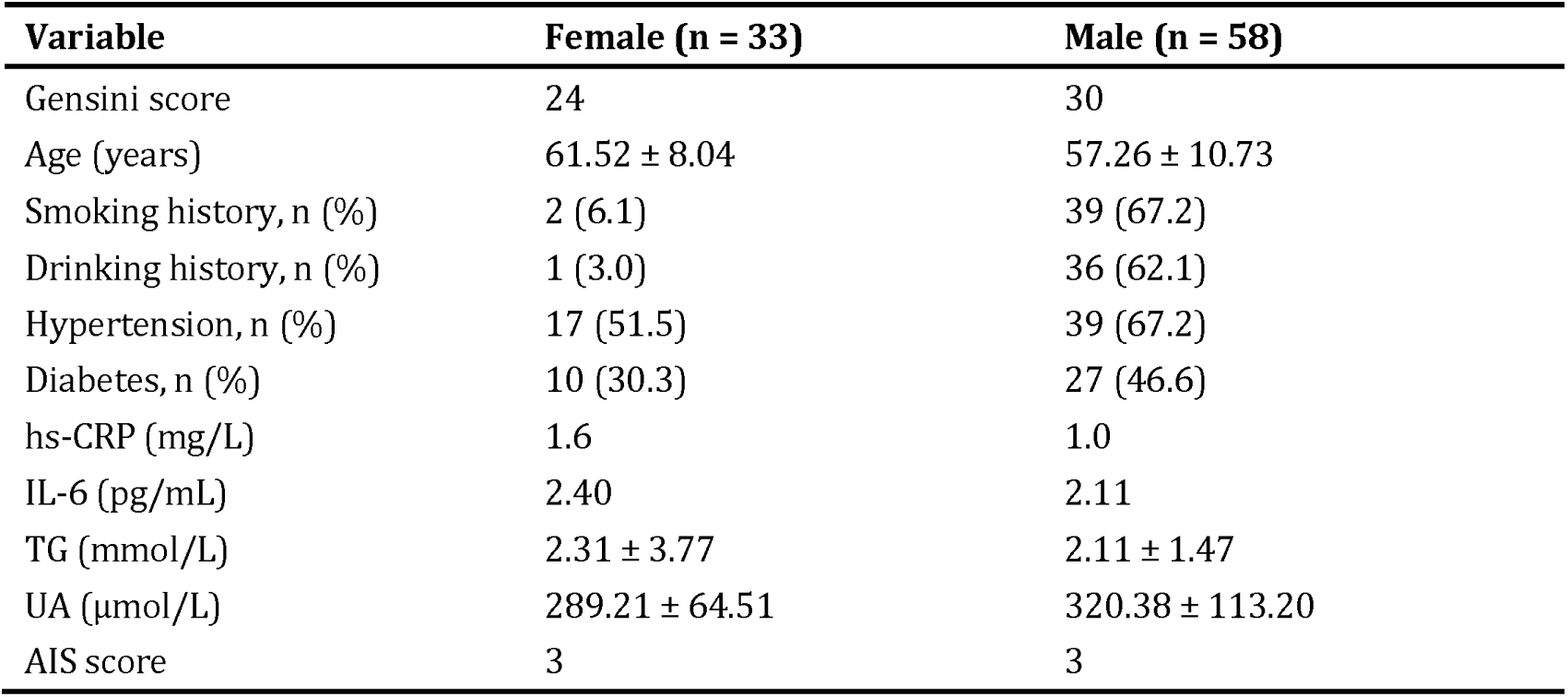
Baseline characteristics of the study population.

### 3.2 Correlation analysis between risk factors and Gensini

To assess the strength of association between individual risk factors and the Gensini score by gender, patients were stratified into female and male groups, and Spearman correlation analysis was performed. The results are presented in Table 2.

**Table 2:** Correlation analysis between risk factors and Gensini score in different genders.

|  | Female |  | Male |  |
| --- | --- | --- | --- | --- |
|  | *r* | *P* | *r* | *P* |
| IL-6 | 0.61 | <0.001* | -0.06 | 0.679 |
| hs-CRP | 0.43 | 0.012* | 0.01 | 0.944 |
| Diabetes | 0.36 | 0.037* | 0.09 | 0.502 |
| SIIRI | 0.29 | 0.107 | 0.22 | 0.104 |
| hypertension | 0.17 | 0.338 | 0.18 | 0.179 |
| AIS | 0.15 | 0.419 | -0.12 | 0.354 |

\* indicates statistically significant

The analysis revealed that, among female patients, the Gensini score was significantly and positively correlated with several inflammatory and metabolic markers. Interleukin-6 (IL-6) showed the strongest correlation (r = 0.61, P < 0.001), followed by high-sensitivity C-reactive protein (hs-CRP) (r = 0.43, P = 0.012) and diabetes (r = 0.36, P = 0.037). These findings suggest that, in women, heightened inflammatory activity and the presence of diabetes are closely linked to greater severity of coronary artery disease.

In contrast, no statistically significant correlations were observed between the examined risk factors and the Gensini score in male patients (all P > 0.05). This significant gender disparity is further illustrated in Figure 1.

**Figure 1.**
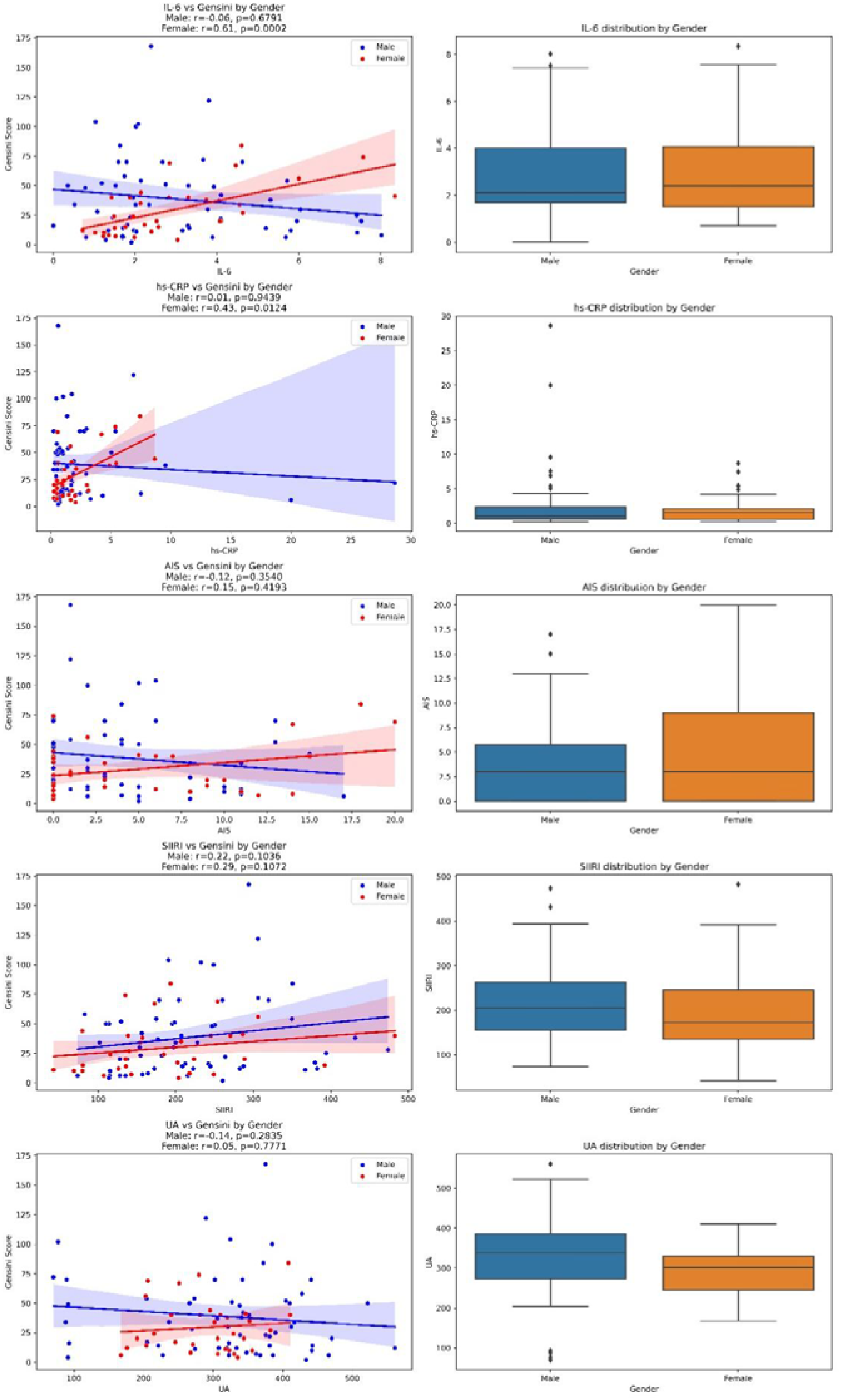
Scatter plot and boxplot of risk factors and Gensini scores in patients of different genders. In female patients, the inflammatory indicators IL-6 and hs-CRP were significantly positively correlated with Gensini, and diabetes was correlated with Gensini; in male patients, the above indicators had basically no correlation with Gensini.

### 3.3 Research on clinical significance and predictive ability

Based on the findings of section 3.2, the Gensini score in female patients with coronary artery disease was closely associated with inflammatory markers and diabetes-related indicators. As a continuous and quantitative measure, the Gensini score reflects both the severity and the extent of coronary atherosclerosis. A higher score indicates a greater disease burden [7]. Previous studies have demonstrated that this scoring system is a strong predictor of major adverse cardiovascular events (MACE) and provides valuable guidance for clinical decision-making, including drug therapy, percutaneous coronary intervention (PCI), and coronary artery bypass grafting [8]. Building upon these findings, we conducted further analyses to establish an evaluation model based on the Gensini score.

#### 3.3.1 Multiple Linear Regression Analysis

We performed multiple linear regression on 33 female patients with coronary artery disease, with the Gensini score as the dependent variable. Independent variables included AIS, hypertension, diabetes, IL-6, hs-CRP, and the interaction term between hs-CRP and diabetes. The model demonstrated a strong overall fit (R² = 0.823, adjusted R² = 0.783, F = 20.19, P < 0.001).

As shown in Table 3, IL-6, hs-CRP, AIS, diabetes, and hypertension were all significantly associated with the severity of coronary artery disease, as reflected by the Gensini score. Notably, the interaction between hs-CRP and diabetes indicates that the effect of inflammatory markers on coronary artery disease is modulated by metabolic status. This finding suggests that the role of hs-CRP is not static but varies depending on whether the patient has diabetes, reflecting a typical moderation effect.

**Table 3:** Multiple linear regression parameters of Gensini score for women.

| Variables | $\beta$ | SE | P | 95% CI |
| --- | --- | --- | --- | --- |
| Intercept | -13.466 | 4.454 | 0.006* | -22.621 ~ -4.311 |
| AIS | 1.223 | 0.300 | <0.001* | 0.608 ~ 1.839 |
| hypertension | 9.369 | 3.974 | 0.026* | 1.201 ~ 17.537 |
| diabetes | 27.968 | 6.112 | <0.001* | 15.404 ~ 40.533 |
| IL-6 | 5.094 | 1.107 | <0.001* | 2.818 ~ 7.370 |
| hs-CRP | 5.802 | 1.006 | <0.001* | 3.734 ~ 7.870 |
| hs-CRP : diabetes | -6.756 | 2.292 | 0.007* | -11.466 ~ -2.046 |

\* indicates statistically significant

#### 3.3.2 Logistic Regression

In clinical practice, patients are often stratified into risk categories according to tertiles or specific cut-off values of the Gensini score [9]. For example, low risk is typically defined as ≤ 20, intermediate risk as 20–50, and high risk as ≥ 50. In this study, based on both clinical experience and data distribution, we used a cut-off of 20: patients with a Gensini score ≤ 20 were classified as the low-risk group, whereas those with a score > 20 were classified as the higher-risk group. A multinomial logistic regression analysis was then performed with the dichotomized Gensini score (GSH) as the dependent variable. Figure 2 and Table 4 demonstrate that IL-6, hs-CRP, diabetes, LDL-TC, and hypertension were all significantly associated with GSH classification.Figure 2 Heatmap of the correlation between risk factors and GSH in female patients.

**Figure 2.**
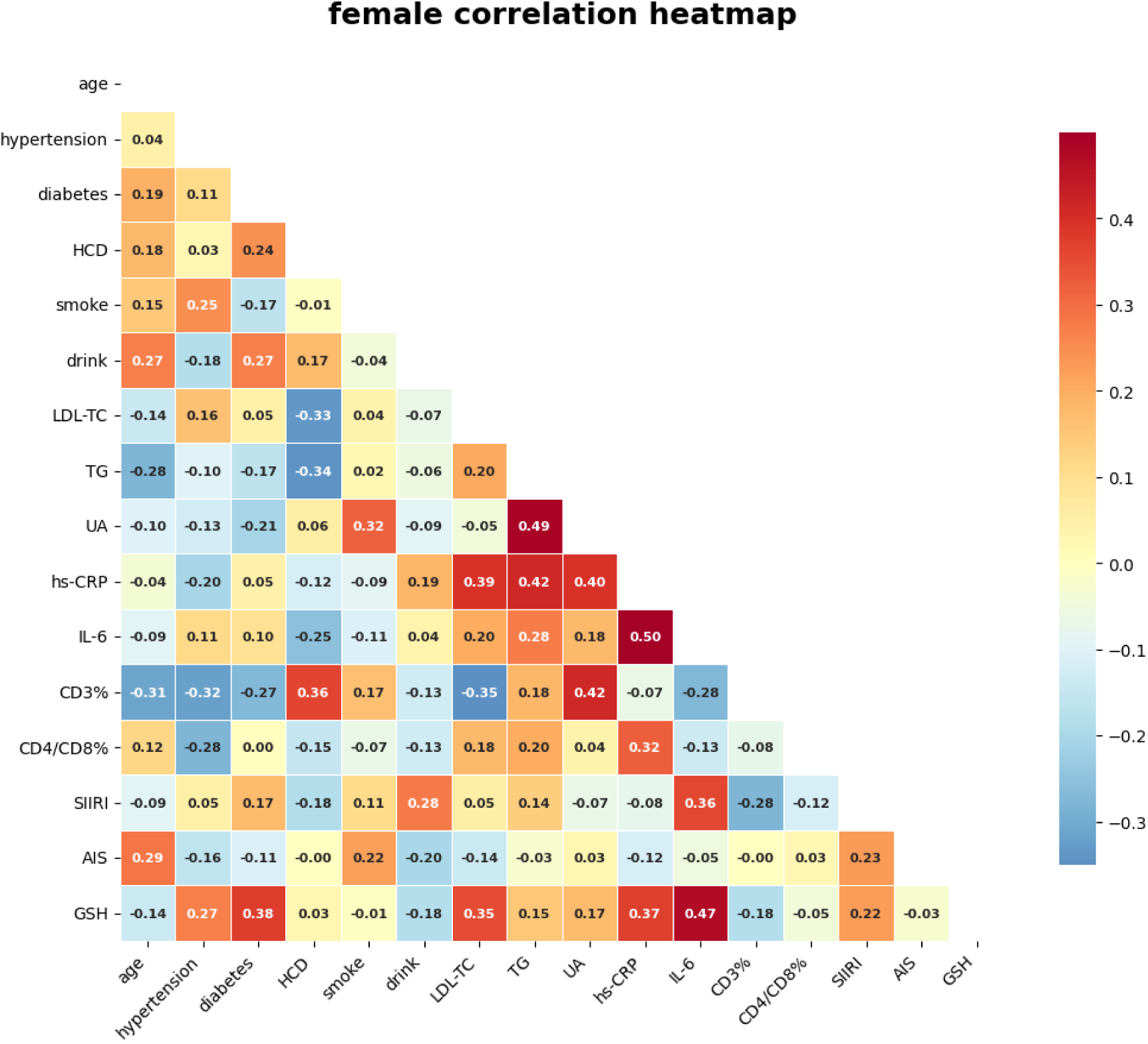
Heatmap of the correlation between risk factors and GSH in female patients.

**Table 4.** Ranking of correlations between risk factors related to GHS.

| variable | r | P |
| --- | --- | --- |
| IL-6 | 0.4712 | 0.0056* |
| diabetes | 0.3758 | 0.0311* |
| hs-CRP | 0.3726 | 0.0327* |
| LDL-TC | 0.3535 | 0.0436* |
| hypertension | 0.2721 | 0.1256 |
| SIIRI | 0.2229 | 0.2125 |
| UA | 0.1719 | 0.3387 |
| CD3% | -0.1815 | 0.3120 |
| AIS | -0.0261 | 0.8853 |

\* indicates statistically significant

In the logistic regression analysis, the dataset was randomly divided into a training set (80%) and a test set (20%). The final model demonstrated satisfactory predictive performance. As shown in Figure 3, the area under the ROC curve (AUC) was 96.4% for the training set and 83.3% for the test set, indicating good model discrimination in both datasets.

**Figure 3.**
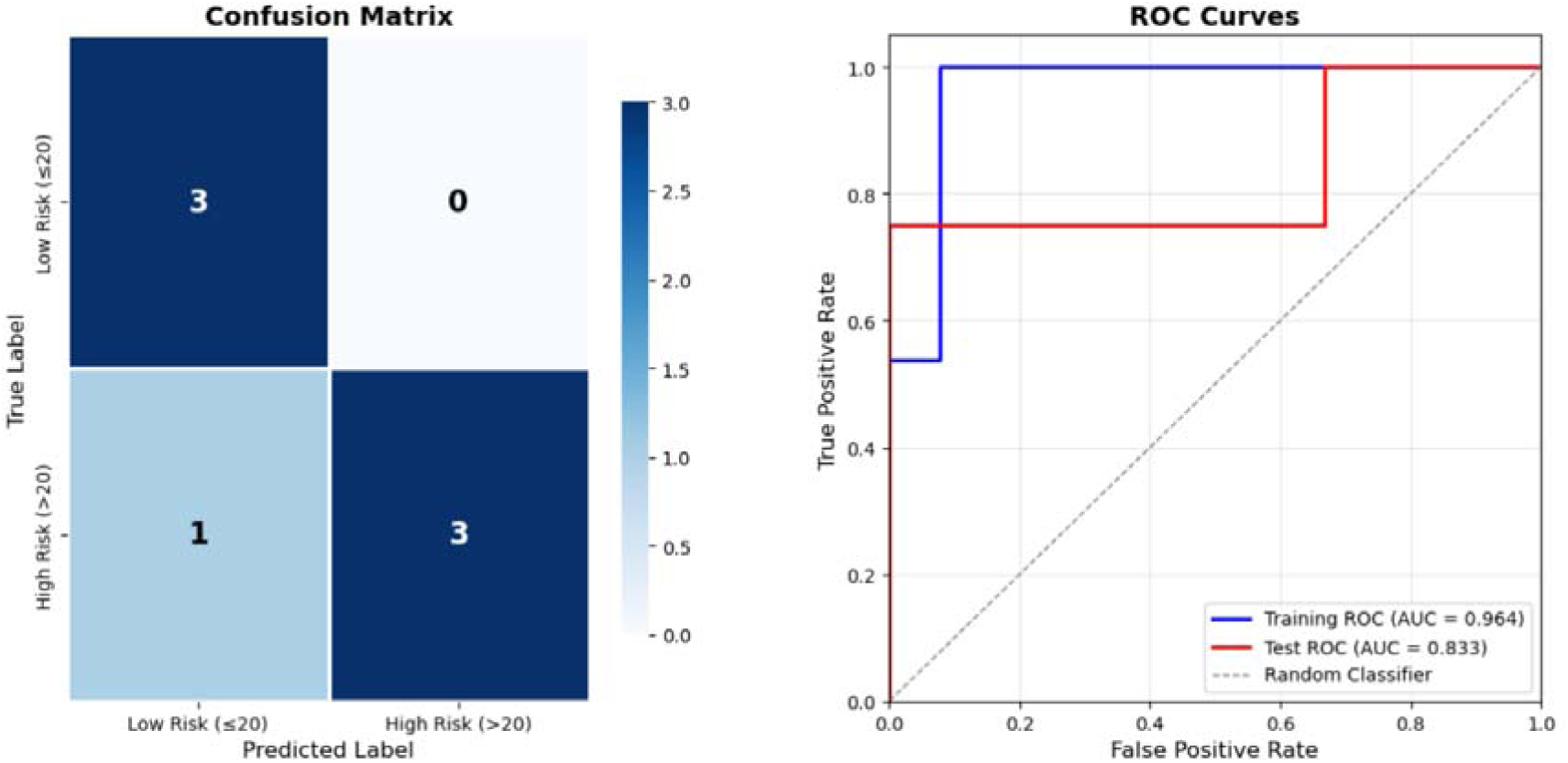
Logistic regression analysis results. Note: The left panel shows the confusion matrix, indicating that among the seven patients in the test set, only one was misclassified as a false negative, while the remaining six were correctly predicted. The right panel presents the receiver operating characteristic (ROC) curve, with the area under the curve (AUC) reaching 96.4% for the training set and 83.3% for the test set.

Figure 4 illustrates the feature importance analysis, which quantifies the relative contribution of each risk factor to the model’s predictive output. The analysis reveals that hypertension, diabetes, hs-CRP, and IL-6 were the most influential predictors. LDL-C and UA also demonstrated a substantial impact on the model’s predictions. In contrast, CD3% and the Athens Insomnia Scale (AIS) score made a comparatively smaller contribution.

**Figure 4.**
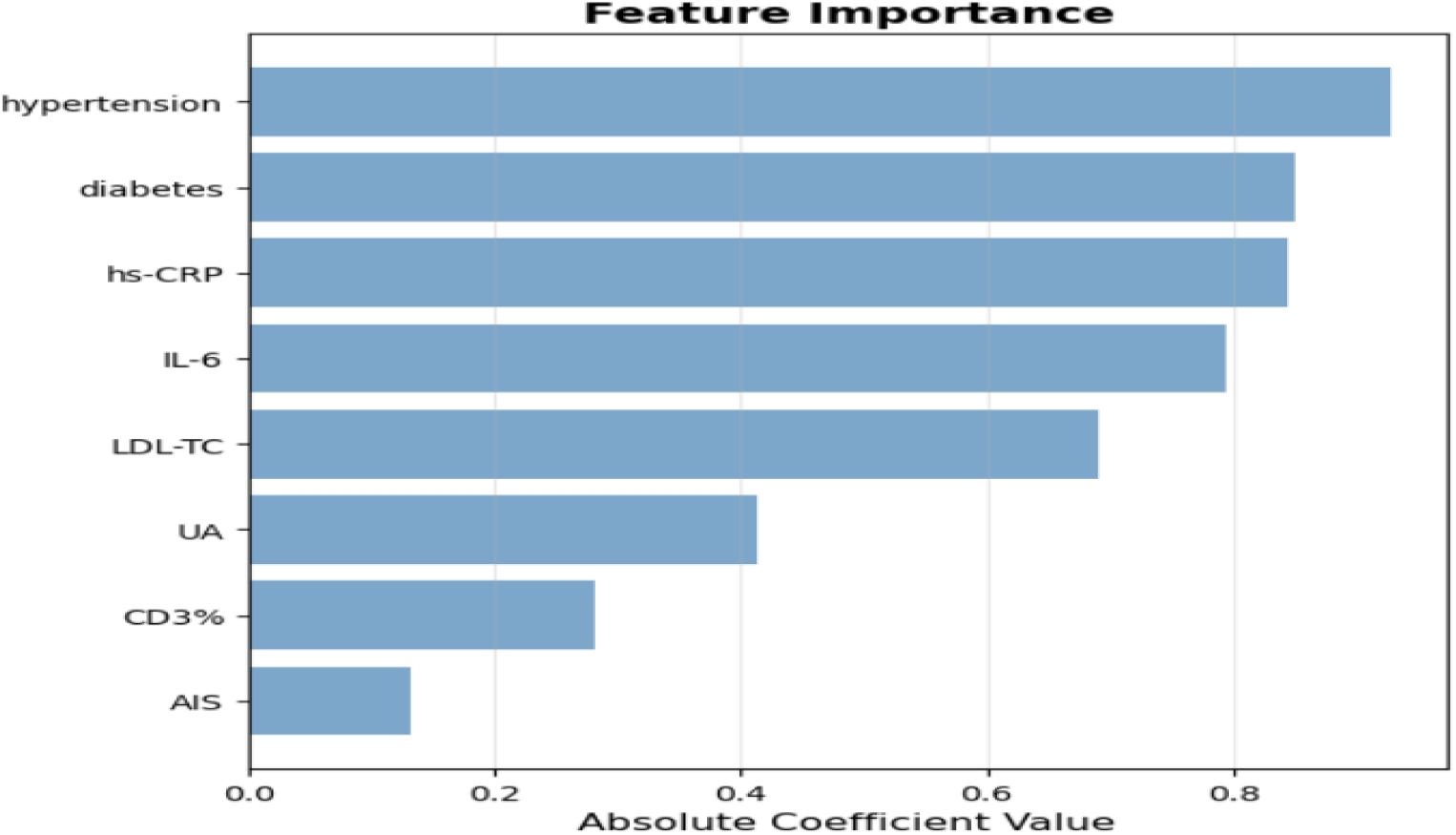
Bar chart of the importance of risk factors in the high-risk group.

The specific coefficients for the final logistic regression model are detailed in Table 5:

**Table 5:** Multinomial logistic regression model parameters.

| hypertension | diabetes | hs-CRP | IL-6 | LDL-TC | UA | CD3% | AIS |
| --- | --- | --- | --- | --- | --- | --- | --- |
| 0.9277 | 0.8511 | 0.8444 | 0.7935 | 0.6912 | 0.4140 | 0.2820 | 0.1307 |

## 4. Discussion

Traditionally, coronary artery disease (CAD) has been attributed to large, focal atherosclerotic plaques causing significant coronary artery narrowing (i.e., obstructive CAD). However, mounting evidence indicates that this pattern is predominantly observed in men.

Women, particularly younger women, often present with a different phenotype, including non-obstructive CAD (NOCAD), diffuse atherosclerosis, and coronary microvascular disease (CMD). In studies of mental stress-induced myocardial ischemia (MSIMI), the occurrence of MSIMI in men was significantly associated with the severity of obstructive CAD, as measured by the Gensini score. In contrast, although women exhibited a higher incidence of MSIMI, no association was observed between MSIMI and the Gensini score [10]. This suggests that in women, myocardial ischemic symptoms and adverse cardiovascular events are frequently driven not by large, flow-limiting plaques, but by functional abnormalities such as endothelial dysfunction and microvascular spasm. These functional alterations may not be adequately captured by the Gensini score, which primarily quantifies anatomical stenosis. Consequently, in women, a high Gensini score may represent only the "tip of the iceberg"—the advanced stage of a prolonged, systemic disease process (e.g., systemic inflammation and metabolic dysregulation) culminating in plaques large enough to be detected and scored on coronary angiography. In men, by contrast, a high Gensini score more directly reflects the primary pathological process of focal plaque growth. Thus, in women, the correlation between systemic markers and the Gensini score may reflect the cumulative and concentrated effects of systemic imbalances driving disease progression.

In this study, multidimensional statistical analyses were conducted to explore the complex factors influencing the Gensini score in female patients with CAD. Univariate linear correlation analyses revealed significant positive associations between the Gensini score and systemic inflammatory markers (IL-6, hs-CRP) as well as diabetes. Multiple linear regression further identified IL-6, hs-CRP, AIS, diabetes, and hypertension as significant contributors to CAD severity, suggesting that unique inflammatory-metabolic processes in women may underpin the development and progression of CAD. Moreover, insomnia may exacerbate systemic inflammation, potentially accelerating atherosclerosis progression. To identify patients at relatively high risk (Gensini score >20), a logistic regression prediction model was constructed. The model demonstrated that, in addition to traditional risk factors such as hypertension and diabetes, inflammatory markers (IL-6, hs-CRP) and metabolic markers, including high- and low-density lipoprotein cholesterol and uric acid, contributed significantly. The model exhibited excellent predictive performance, with areas under the curve (AUCs) of 96.4% and 83.3% in the training and test sets, respectively. These findings indicate that an assessment model integrating multidimensional risk factors can reliably identify high-risk female patients with CAD, providing robust support for clinicians in developing early and precise intervention strategies.

Inflammation is a central driver of the initiation and progression of atherosclerosis. In this study, inflammation-related markers (IL-6, hs-CRP) exhibited stronger correlations in women, highlighting a sex-specific inflammatory response pattern. Women appear to have a higher baseline inflammatory potential, which is closely linked to their cardiovascular risk. Previous studies[11] have shown that, even after adjusting for traditional cardiovascular risk factors, women generally exhibit higher hs-CRP levels than men across various racial subgroups, suggesting a physiological baseline of chronic low-grade inflammation. Moreover, multiple studies have confirmed a positive association between hs-CRP levels and Gensini scores, indicating that higher systemic inflammation corresponds to more severe coronary atherosclerosis [12]. Sex differences in inflammatory responses under stress are particularly striking. In patients with stable CAD, women exhibited more intense inflammatory responses to mental stress than men, which translated directly into clinical risk. Specifically, in women, each standard deviation increase in stress-induced IL-6 was associated with a 56% higher risk of future major adverse cardiovascular events (MACE) (HR: 1.56), whereas no such association was observed in men [13]. These findings suggest that women’s immune systems are not only more active at rest (higher baseline hs-CRP) but also more reactive to acute stimuli (e.g., mental stress or tissue injury), as reflected by heightened IL-6 responses. This heightened inflammatory state provides a mechanistic link to endothelial injury, plaque instability, and ultimately higher Gensini scores. Accordingly, the strong correlations observed in this study between IL-6 (r = 0.61, P < 0.001) and hs-CRP (r = 0.43, P = 0.012) in women are well supported by existing evidence.

In high-risk female populations, traditional risk factors such as hypertension, diabetes, and elevated LDL-C have been shown to affect Gensini scores, while inflammatory markers (IL-6, hs-CRP) maintain a strong correlation, indicating that the severity of coronary lesions in women is more strongly driven by inflammatory-metabolic factors. Notably, our study identified a significant interaction between diabetes and hs-CRP in multiple linear regression (β = –6.756). This implies that the effect of hs-CRP on the Gensini score varies depending on diabetic status: in non-diabetic patients (diabetes = 0), the effect of hs-CRP corresponds to its main effect (+5.802), whereas in diabetic patients (diabetes = 1), the effect is the sum of the main and interaction effects, i.e., 5.802 + (–6.756) = –0.954. In other words, in diabetic patients, higher hs-CRP may slightly reduce the Gensini score. This phenomenon may relate to the chronic inflammatory milieu characteristic of diabetes [14]. Diabetic patients often exhibit a combination of hyperglycemia, dyslipidemia (high triglycerides, low HDL-C), hypertension, insulin resistance, and oxidative stress. In this high-risk context, the predictive value of hs-CRP as a single biomarker is diminished, weakening its ability to explain the severity of coronary atherosclerosis. In essence, elevated baseline inflammation in diabetes attenuates the marginal effect of hs-CRP. This observation suggests that diabetes may partially “mask” the relationship between hs-CRP and CAD severity, underscoring the central role of diabetes itself as a driver of systemic inflammation. Supporting this, a study of coronary artery calcification (CAC) found that in women with diabetes, the association between elevated CRP levels and CAC was not statistically significant (OR = 2.6, P = 0.07), whereas in women without diabetes, the association remained significant (OR = 3.1, P = 0.04), though attenuated after adjusting for body mass index [15]. Similarly, in patients with acute myocardial infarction, the HR for MACE in non-diabetic women was 1.39 (1.14–1.85, P = 0.002), compared with 1.02 (0.73–1.23, P = 0.453) in diabetic women [16]. These findings are consistent with our results, indicating that the predictive power of hs-CRP for cardiovascular events is consistently weakened or eliminated in the presence of diabetes.

Insomnia is not only a "behavioral" risk factor for CAD, but also a direct biological damage. The sleep index (AIS score) in this study showed a strong correlation with the Gensini score in female patients in the multiple linear regression analysis (see Table 3, β = 1.223, p < 0.001), revealing a pathological pathway from the brain to the heart that is particularly unobstructed in women. Insomnia leads to blood pressure fluctuations, increased heart rate and oxidative stress response through sympathetic overactivation (such as increased norepinephrine) and hypothalamic-pituitary-adrenal axis (HPA axis) disorders, promoting metabolic dysfunction (such as insulin resistance), exacerbating inflammatory response, and directly increasing cardiovascular risk [17]. This over-activation state then leads to endothelial dysfunction (manifested as decreased flow-mediated dilation (FMD) [18]) and systemic inflammation (manifested as increased WBC [19] and CRP [20]). Endothelial dysfunction and inflammation are the two core initiators of atherosclerosis [21]. The long-term accumulation of this atherosclerotic process ultimately results in the formation of coronary artery plaques, and the extent and severity of these plaques are precisely quantified by the Gensini score. This clear pathophysiological pathway provides a strong mechanistic argument for the association between AIS and the Gensini score, and also makes the current lack of direct research particularly prominent.

Interestingly, the correlation between the AIS and the Gensini score was not significant in univariate correlation analysis. However, in multiple regression analysis, when important factors such as diabetes, hypertension, and inflammatory markers were included in the model, the effect of the AIS on the Gensini score became clear and highly significant. The AIS was significantly correlated with the Gensini score, with a one-point increase in the AIS corresponding to a 1.223-point increase in the Gensini score. This finding suggests that when studying disease-related factors, relying solely on a single correlation analysis between the independent and dependent variables may overlook potential confounding or interactions. Only through a multidimensional, comprehensive analysis can the true role of each indicator be more comprehensively and accurately revealed.

The role of uric acid in CAD, whether as a biomarker or a causal factor, has been widely debated [22]. Hyperuricemia can contribute to the pathogenesis of atherosclerosis, hypertension, and heart failure through mechanisms involving oxidative stress, inflammatory responses, and endothelial dysfunction. Thus, uric acid is considered not only a marker of cardiovascular disease but also a potential therapeutic target. In women, higher serum uric acid (SUA) levels have been significantly associated with a greater prevalence of severe CAD (p < 0.001; adjusted OR [95% CI] = 1.29 [1.03–1.62], p = 0.03) [23]. Consistently, in our Gensini score logistic regression analysis, uric acid contributed modestly to the prediction model, suggesting that even moderately elevated uric acid levels warrant clinical attention in female patients (mean SUA in this study: 289 μmol/L).

CD3⁺ T cells are critical effector cells within the immune system, central to both immune regulation and inflammatory responses. The metric CD3%, which represents the proportion of CD3⁺ T lymphocytes within the total peripheral lymphocyte population, serves as a common "pan-T cell marker." As such, it primarily reflects the total T cell count. However, the pathogenesis of atherosclerosis is principally mediated by T cell-driven immune responses to specific antigens, such as oxidized low-density lipoprotein (oxLDL). Crucially, this process is dependent on the function of the T cell receptor (TCR), not the CD3 molecule itself. This distinction underscores a fundamental limitation of the CD3% indicator: while it quantifies T cell numbers, it fails to provide insight into their functional polarity, activation status, or antigen specificity—characteristics that ultimately determine their pro-atherogenic or protective roles. This theoretical limitation aligns with our finding that CD3% demonstrated a weak association with the Gensini score.

Indicators derived from routine complete blood counts, such as the neutrophil-to-lymphocyte ratio (NLR) and lymphocyte-to-monocyte ratio (LMR), have emerged as robust predictors of coronary artery disease (CAD) prognosis. A substantial body of evidence shows that an elevated NLR, reflecting relative neutrophilia and lymphopenia, and a reduced LMR are strongly associated with increased atherosclerotic burden, as quantified by angiographic metrics such as the Gensini score, and with adverse clinical outcomes (MACE). In this study, we evaluated the systemic immune-inflammation index (SIIRI), a composite marker integrating neutrophils and monocytes—key components of innate immunity—with lymphocytes, which are central to adaptive immunity, to provide a more holistic assessment of systemic immune-inflammatory status. Contrary to expectations, our results indicated a weak association between SIIRI and the study endpoints, both in the overall cohort and within the female subgroup. This finding suggests that, while composite indices like SIIRI may offer a macroscopic view of systemic inflammation, the pathophysiology of vascular damage cannot be fully captured by any single inflammatory marker. We propose that vascular damage represents a complex pathological outcome arising from the multifactorial interplay of inflammation with coexisting metabolic and endocrine dysregulation.

## 5. Study Limitations

This study was a cross-sectional investigation with a relatively small sample size. It focused on selected factors influencing the Gensini score in female patients with coronary artery disease and did not include longitudinal follow-up of the relatively high-risk group (Gensini score >20). Consequently, the findings have inherent limitations in terms of causal inference and generalizability. Future studies should expand the sample size and incorporate prospective follow-up, with particular attention to major adverse cardiovascular events (MACE), to further elucidate risk factors affecting prognosis in high-risk patients and to provide evidence-based guidance for clinical decision-making, including medical therapy, percutaneous coronary intervention (PCI), or coronary artery bypass grafting (CABG).

Additionally, this study identified a significant interaction between diabetes and hs-CRP, suggesting that diabetes may modulate the relationship between systemic inflammation and CAD progression. In future research, we plan to integrate additional metabolic markers, such as glycated hemoglobin (HbA1c), triglycerides (TG), and high-density lipoprotein cholesterol (HDL-C), to construct a multidimensional interactive risk model. This approach aims to enable a more accurate and personalized assessment of cardiovascular risk in patients with diabetes.

## 6. Conclusion

In conclusion, the severity of coronary atherosclerosis in female patients is driven by a complex interplay of inflammatory, metabolic, and lifestyle-related factors, exhibiting risk patterns distinct from those observed in men. This underscores the necessity of a sex-specific, multidimensional approach to risk stratification in this population, moving beyond traditional risk paradigms. The predictive models developed herein, incorporating variables such as hypertension, diabetes, hs-CRP, IL-6, LDL-C, uric acid, and a history of AIS, demonstrated substantial explanatory power (R² = 0.823 for the linear regression) and excellent discriminatory performance in identifying high-risk individuals (AUC = 0.833 for the logistic regression in the test set). Notably, our identification of a significant interaction between hs-CRP and diabetes highlights the intricate and context-dependent nature of these risk factors. The integration of broader clinical datasets in future studies may yield more robust and reliable models, providing stronger support for personalized risk stratification and evidence-based management in women with CAD.

## Data Availability

All data produced in the present study are available upon reasonable request to the authors

## Conflicts of Interest

There are no conflicts of interest in this study.

## Author Contributions

Conceptualization, Guoxia An; methodology, Xiaohua Liu,Guoxia An; software, Xiaohua Liu, Ruizhe Li; Validation, Xiaohua Liu,Guoxia A,Ruizhe Li; investigation, Guoxia An, Weimin Guo; Data acquisition, Guoxia An, Weimin Guo; Data analysis& exploration : Xiaohua Liu, Ruizhe Li

## Data Availability

The datasets used and/or analyzed during the current study are available from the corresponding author upon reasonable request.

## Acknowledgments

The authors would like to thank all participants and staff of the Second Hospital of Shanxi Medical University for their contributions to this study.

## Funding

This research did not receive any specific grant from funding agencies in the public, commercial, or not-for-profit sectors.

## Notes

### Competing Interest Statement

The authors have declared no competing interest.

### Funding Statement

This study did not receive any funding

### Author Declarations

This study was approved by the Institutional Review Board of Second Hospital of Shanxi Medical University. As the study employed a retrospective design using medical records, the requirement for informed consent was waived by the IRB.All data were anonymized prior to analysis

### Summary of Updates

received many spam emails, so removed sensitive personal information of Author.

